# Revision Behavior and Explainability in Adaptive LLM Swarms for ICU Mortality Risk Prediction: A Two-Dataset Evaluation

**DOI:** 10.64898/2026.09.01.26361960

**Authors:** Callum Anderson

## Abstract

**Purpose:** To evaluate how adaptive LLM swarm revision changes ICU mortality-risk outputs and to characterize evidence use, explanation indicators, auditability, and computational burden in final adaptive outputs relative to an independently executed fixed-voting (FV) architecture.

**Methods:** We retrospectively analyzed 1,607 eICU encounters (1,500 stays) and 1,607 ICU-2012 encounters. Initial (ASI) and final (ASF) adaptive outputs were compared within runs for revision engagement and risk-score drift; final ASF and separately generated FV outputs were compared for evidence use, explanation indicators, auditability, and computation. Paired differences and 95% confidence intervals used 10,000 hospital-stay-clustered bootstrap replicates.

**Results:** At least one specialist revision trace occurred in 84.32% of eICU and 56.44% of ICU-2012 encounters. Mean ASF-minus-ASI risk-score changes were **+0.0810** and **+0.0539**; 757 of 761 0.50-threshold crossings moved toward mortality, without clear AUROC or AUPRC improvement. In the independent benchmark, ASF explanations contained 1.26 and 0.55 more supporting-evidence items than FV, but counterevidence acknowledgement was 21.59 and 7.47 percentage points lower and unsupported-claim flags were 1.43 and 0.68 points higher. All final records met the reconstruction-completeness criterion, although ASF generated more warnings and required 1.97 and 1.52 times the FV runtime.

**Conclusion:** Adaptive revision materially changed swarm operating behaviour. Independently, final ASF outputs showed greater supporting-evidence use but less balanced evidence engagement, more process warnings, and greater computational burden than FV. These automated artifact-level findings do not establish superior explanation quality or isolate revision as their cause.

## 1 Introduction

Large language models (LLMs) are increasingly being evaluated for evidence synthesis and clinical decision support, although their performance varies with the target task, prompting strategy, and model architecture [8, 18, 27]. Multi-agent systems distribute tasks across LLM instances whose outputs can be combined, challenged, or revised, thereby approximating human clinical team dynamics. Role-differentiated and LLM-driven multi-agent systems have reported gains across medical decision-making, diagnostic benchmarks, and EHR-based mortality and readmission prediction [4, 14, 15, 37].

These reported gains do not establish a general benefit. Multi-agent debate may fail to outperform simpler aggregation and can converge on shared misconceptions [7, 31]. Clinical outputs remain vulnerable to contradictions, omissions, and hallucinations [13, 18], yet evaluation is often narrow: only 5% of 519 healthcare LLM studies used real patient-care data, and 95.4% primarily assessed accuracy [3]. Revision may instead increase sensitivity by classifying mortality more often without improving risk-score ranking. Evaluation should therefore distinguish discrimination, calibration, threshold-dependent classification, and clinical utility [28, 34]. Agent roles and evidence routing can also alter class-wise errors under fixed inference conditions [1].

Observable LLM-generated reasoning is not necessarily faithful. A plausible explanation accompanying an individual patient’s prediction does not establish that the model relied on the stated evidence or that the prediction is correct [11], while chain-of-thought may rationalize biased outputs [33] or have limited causal influence on final answers [22]. Externalizing specialist assessments, disagreements, and revisions can create reviewable artifacts, but such artifacts are not evidence of correctness [11, 36]. We therefore assess these traces as auditable process records, focusing on evidence balance, unsupported claims, decision-path reconstructability, integrity warnings, and computational burden.

These questions are especially relevant in intensive care, where prediction must integrate sparse, irregular, and rapidly evolving EHR data. Longitudinal models have demonstrated the feasibility of externally validated dynamic prediction [6, 39]. We evaluated adaptive LLM swarm deliberation on two ICU datasets, quantifying within-run revision effects and benchmarking final ASF against an independently executed FV architecture. Outcomes included score drift, evidence use, explanation indicators, reconstructability, warnings, predictive performance, and computational burden. This was a methodological evaluation rather than a deployment study; no clinical end-user, position in a care pathway, autonomous role, or decision-support use was specified.

## 2 Methods

### 2.1 Study Design and Data Sources

This retrospective cross-dataset study used two comparisons. ASI and ASF were compared within each adaptive execution to assess revision. Separately, final ASF was benchmarked against an independently executed FV architecture. Because ASF and FV did not share specialist calls, their differences represent complete architecture outputs rather than isolated revision effects. Predictive performance was contextual, and scores were treated as uncalibrated.

We analyzed 1,607 ICU encounters from each of two EHR-derived datasets: the eICU Collaborative Research Database Demo, version 2.0.1, derived from eICU-CRD version 2.0 [23, 25], and the PhysioNet/Computing in Cardiology Challenge 2012 dataset, version 1.0.0 (ICU-2012) [30]. Both datasets were obtained through PhysioNet [24]. eICU-CRD contains encounters collected in 2014–2015; record-level calendar dates are not provided in the de-identified ICU-2012 release. Model inputs were structured fields rather than a natural-language evaluation corpus. Analyses were conducted separately by dataset; cross-dataset comparisons were descriptive because of differences in mortality prevalence, clustering, available clinical domains, and specialist participation.

### 2.2 Input Data Preprocessing

For eICU, 2,520 source encounters were screened. Encounters were eligible when hospital-discharge status was available and at least 24 hours of ICU data were recorded (`unitdischargeoffset` ≥ 1,440 minutes). No age or index-encounter restriction was applied, and all 1,607 eligible encounters were retained without further sampling. The final eICU cohort included 141 deaths and 1,466 survivors, corresponding to 8.77% mortality.

For ICU-2012, 12,000 records remained after stays shorter than 48 hours were excluded. Proportional stratified random sampling by mortality selected 1,607 records to match the eICU cohort size and nominal inference workload while preserving the source outcome distribution. The final ICU-2012 sample included 229 deaths and 1,378 survivors, corresponding to 14.25% mortality, compared with 14.23% in the source data (1,707/12,000).

The prediction unit was an ICU encounter. In eICU, 1,607 encounters mapped to 1,500 hospital stays using `patientunitstayid` and `patienthealthsystemstayid`; 94 stays contained repeated encounters, producing 107 additional encounters, with a maximum of three per stay. Encounters within a stay shared the hospital-discharge outcome. ICU-2012 `RecordID` values were used as encounter and clustering identifiers, although they did not establish longitudinal patient linkage. The outcome was all-cause in-hospital mortality, defined by `hospitaldischargestatus = Expired` in eICU and `In-hospital_death = 1` in ICU-2012. For both datasets, model inputs were limited to the first 24 hours of each ICU encounter. Predictions were interpreted at the end of this observation window and targeted subsequent death before hospital discharge.

eICU inputs included demographics, admission details, medical history, diagnoses, laboratory results, and selected medications, infusions, and major treatments. Continuously recorded bedside vital signs were not used; repeated or differently named records of the same item were combined, and treatment data were limited to categories chosen before analysis for their relevance to mortality risk.

ICU-2012 inputs comprised admission descriptors and available physiological and laboratory series, including vital signs, blood gases, Glasgow Coma Scale, ventilation, urine output, weight, and laboratory tests, which were jointly supplied to the laboratory specialist.

Numeric series were summarized by count, first and last values, minimum, maximum, unit, and directional trend. Missing or nonnumeric values were omitted without imputation; absent information was treated as unknown rather than normal. Variable and unit labels were made consistent (e.g., blood urea nitrogen as `BUN` and milligrams per decilitre as `mg/dL`); ICU-2012 FiO_2_ fractions were converted to percentages, whereas eICU measurements otherwise retained their source units.

After unit harmonization, nonmissing numeric observations were retained without physiological plausibility filtering, clipping, or winsorization. This intentionally preserved artifacts such as zero blood pressures and negative temperatures so that system handling of supplied EHR evidence remained observable rather than being optimized through input cleaning. No post hoc correction altered agent inputs or outputs.

For both datasets, each encounter was converted to the same fixed-format JSON structure, divided into clinical domains, and assigned reproducible evidence identifiers; the available content within those domains differed as described above. For example, a shortened laboratory entry could appear as `{“creatinine”:{“evidence_id”: “LAB-CREATININE”, “value”:{“first”:3. 6,”last”:3.5, “unit”:”mg/dL”}}}`. In both datasets, specialists received only their assigned domains and were instructed to cite supplied in-domain evidence, treat missing data as unknown, and avoid chain-of-thought disclosure. Outcome labels were stored separately and joined only during evaluation. Model inputs excluded outcome, discharge, post-window, and provenance fields.

Because ICU-2012 provided no history, diagnosis, medication, infusion, or treatment evidence, two specialists returned `no_domain_evidence` for every encounter in both architectures and were excluded from aggregation, leaving at most two active specialists. These systematic abstentions were distinguished from occasional invalid-output or invalid-citation abstentions when interpreting revision, audit, and computational measures.

### 2.3 Adaptive-Swarm and Fixed-Voting Architectures

Both architectures used the same four domain specialists covering admission and demographics, medical history and diagnoses, laboratory results, and medications, infusions, and treatments. Evidence identifiers encoded domain provenance using `DEM-` and `ADM-` for demographics and admission, `HIS-` and `DX-` for medical history and diagnoses, `LAB-` for laboratory results, and `MED-`, `INF-`, and `TRT-` for medications, infusions, and treatments, respectively.

During the initial assessment for each encounter, the specialists processed their assigned first-24-hour EHR evidence concurrently and remained blinded to the other domains. Each returned a structured assessment containing a continuous mortality risk score on the 0–1 scale, the corresponding threshold-derived binary prediction, and cited evidence identifiers. The domain prefixes defined enforceable evidence boundaries. Specialists could access and cite raw evidence only from their assigned domains, and output validation removed unknown or out-of-domain evidence identifiers. Only validated, non-abstaining assessments contributed to aggregation.

In FV, specialists completed one assessment without communication or revision. The encounter-level score was their unweighted mean, whereas the stored architecture decision used majority vote with the mean score resolving ties. FV generated its own specialist assessments in a separate execution.

The adaptive-swarm architecture (Fig. 1) used the same roles, validation checks, and aggregation rule but generated its own initial assessments. Their saved aggregate formed ASI without additional calls. A deterministic controller triggered revision when votes were tied, scores were excessively dispersed, or high-confidence votes opposed one another.

**Fig. 1.**
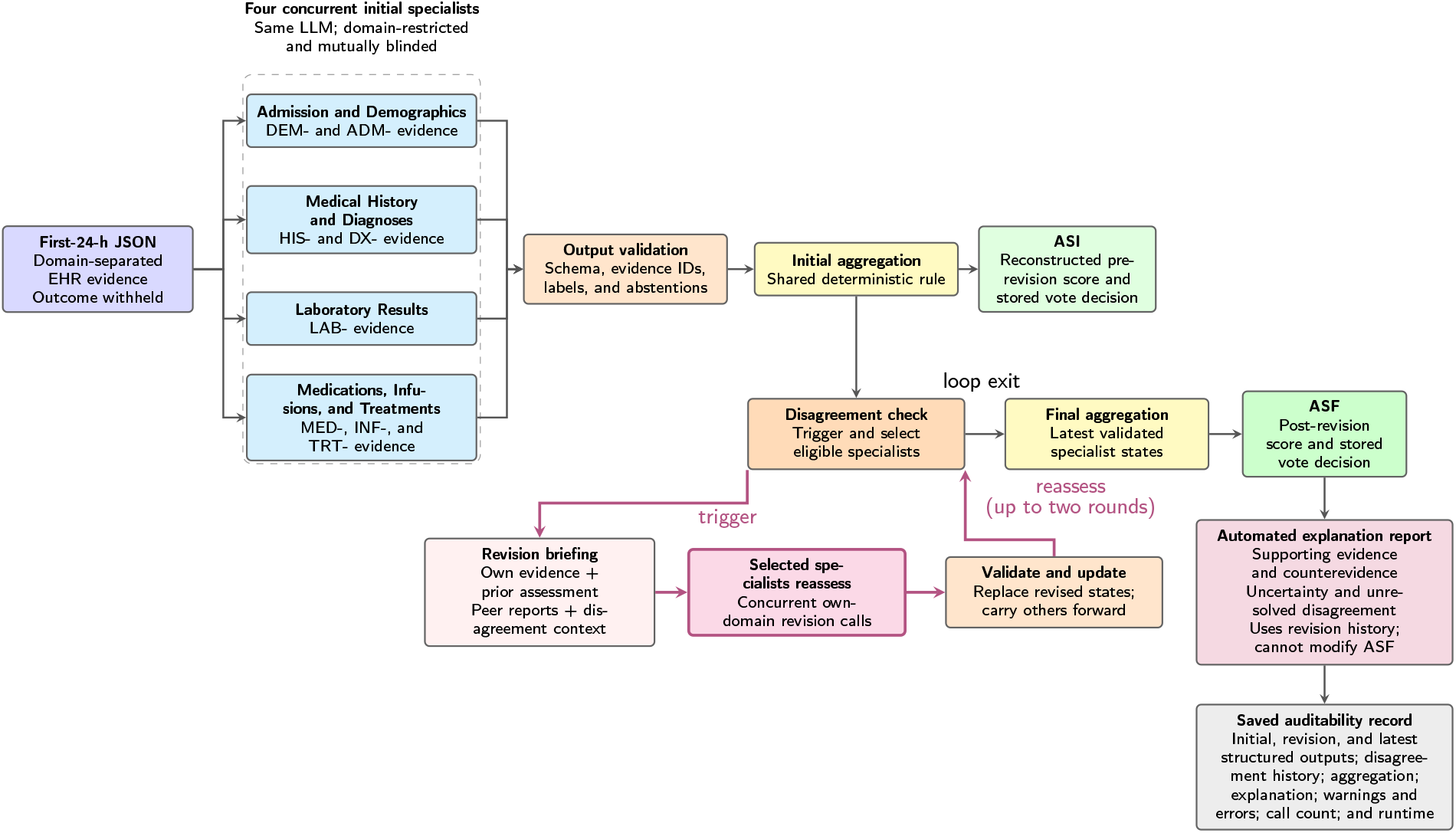
Adaptive-swarm architecture and revision workflow.

When the controller triggered a revision round, the selected specialists were prompted in parallel to reassess their own domain evidence and previous assessment using the identified disagreement and the other specialists’ structured reports. These peer reports included mortality risk scores, binary predictions, confidence, rationales, and cited evidence identifiers, but not the underlying raw EHR evidence from the peers’ domains. Cross-domain information could therefore inform reconsideration only as another specialist’s reported assessment, and revising specialists remained restricted to citing evidence from their own domains. Each selected specialist could retain or revise its assessment. Validated revisions replaced the corresponding specialist states, while other states were carried forward unchanged. The controller performed up to two revision rounds, stopping earlier if the disagreement cleared or no further eligible revision was available. After revision stopped, the latest validated specialist assessments were aggregated using the same rule; this post-revision aggregate was the adaptive-swarm final (ASF) output.

The aggregate score and stored majority-vote decision were distinct fields. All reported threshold-dependent metrics instead used a consistent analytic classification: aggregate mean mortality risk score ≥ 0.50 for ASI, ASF, and FV.

#### Implementation details

All LLM calls used the Q4_K_M-quantized Qwen3.5 27B Ollama model (`qwen3.5:27b-q4_K_M`) [26], served through Ollama’s HTTP `/api/generate` endpoint. Calls used temperature 0, disabled model thinking, non-streaming generation, and JSON-formatted outputs. The model was released in February 2026; its developers did not report the last date of training. Prompts were author-written from the prespecified clinical-domain boundaries, structured-output schemas, and validation rules and were not tuned against evaluation outcome labels. The workflow, controller, and agent-state transitions were implemented in Python 3.13 using LangGraph. Both architectures shared the model, inference settings, specialist roles and schemas, validation rules, and deterministic aggregation procedure. They were executed independently; specialist responses were not reused across architectures. ASF additionally used revision prompts and supplied revision history to its final explanation prompt. Accordingly, ASF–FV comparisons include independently generated specialist outputs and architecture-specific explanation contexts. Prompts, configurations, and software dependencies are available in the experiment repository.

### 2.4 Evaluation Measures

Revision behavior was the primary evaluation. Within adaptive runs, ASF was compared with ASI for score changes, 0.50 threshold crossings, and predictive changes. Scores were treated as uncalibrated, with mortality positive. AUROC and AUPRC measured discrimination, and Brier score measured prediction error. Threshold-dependent metrics used aggregate mean score ≥0.50, not the stored majority-vote decision. Metrics included sensitivity, specificity, predictive values, balanced accuracy, Matthews correlation coefficient, F1, score-threshold mortality classification prevalence, and secondary accuracy.

Evidence-use and automated explanation indicators included citation validity, specialist-evidence coverage, evidence counts, counterevidence acknowledgement, uncertainty disclosure, unsupported-claim flags, contradictions, and validation failures. Rates used one final explanation per encounter. ASF–FV contrasts compared independent end-to-end executions, not revision alone, and were not human-validated measures of explanation quality.

Process auditability included reconstruction-completeness, revision traces, warnings, fallbacks, and abstentions. Completeness required the latest specialist outputs, aggregation rule, final explanation object, and audit-warning field; it measured artifact availability, not correctness. No composite auditability score was calculated.

Computational burden comprised model calls and wall-clock runtime per encounter.

### 2.5 Statistical Analysis

Analyses were separate by dataset. Paired hospital-stay-clustered bootstrap resampling retained all encounters within sampled stays and applied identical encounter samples to ASI, ASF, and FV. Each of 10,000 replicates (seed 20260801) sampled 1,500 eICU or 1,607 ICU-2012 clusters; ICU-2012 clustering was equivalent to encounter resampling. Percentile 95% confidence intervals used the 2.5th and 97.5th percentiles, and the analytic threshold was fixed at 0.50. ASF–FV pairing was by encounter, not shared model call; intervals therefore capture cohort-sampling uncertainty conditional on the two observed runs, not between-run LLM variability. No multiplicity adjustment was applied. Model calls and runtime were descriptive. Analyses used Python 3.13. Reporting was informed by the TRIPOD+AI and TRIPOD-LLM frameworks [5, 9].

## 3 Results

### 3.1 Cohort Characteristics

Both cohorts contained 1,607 ICU encounters; eICU included repeated encounters within hospital stays and had lower mortality prevalence than ICU-2012 (8.77% versus 14.25%; Table 1).

**Table 1.**
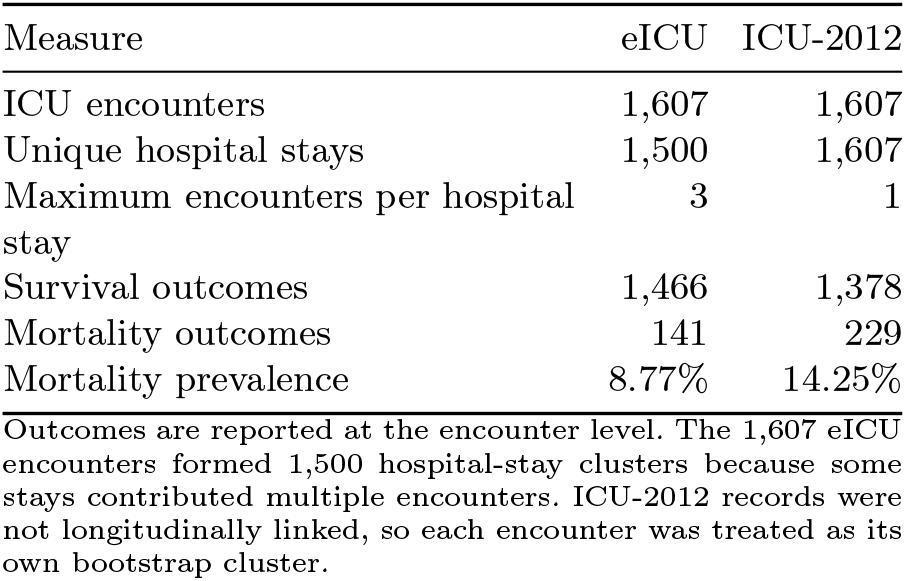
Cohort structure and outcome distribution.

### 3.2 Revision-Associated Mortality Risk Score Drift

At least one revision trace was recorded for 1,355 eICU encounters (84.32%) and 907 ICU-2012 encounters (56.44%) (Table 2). Scores increased more often than they decreased. Of 761 crossings of the 0.50 analytic threshold, 757 (99.47%) moved toward the at-or-above-threshold mortality classification and four (0.53%) moved oppositely. These score-derived classes are distinct from the stored majority-vote decisions. Figure 2 shows the resulting directional operating-point shift.

**Table 2.**
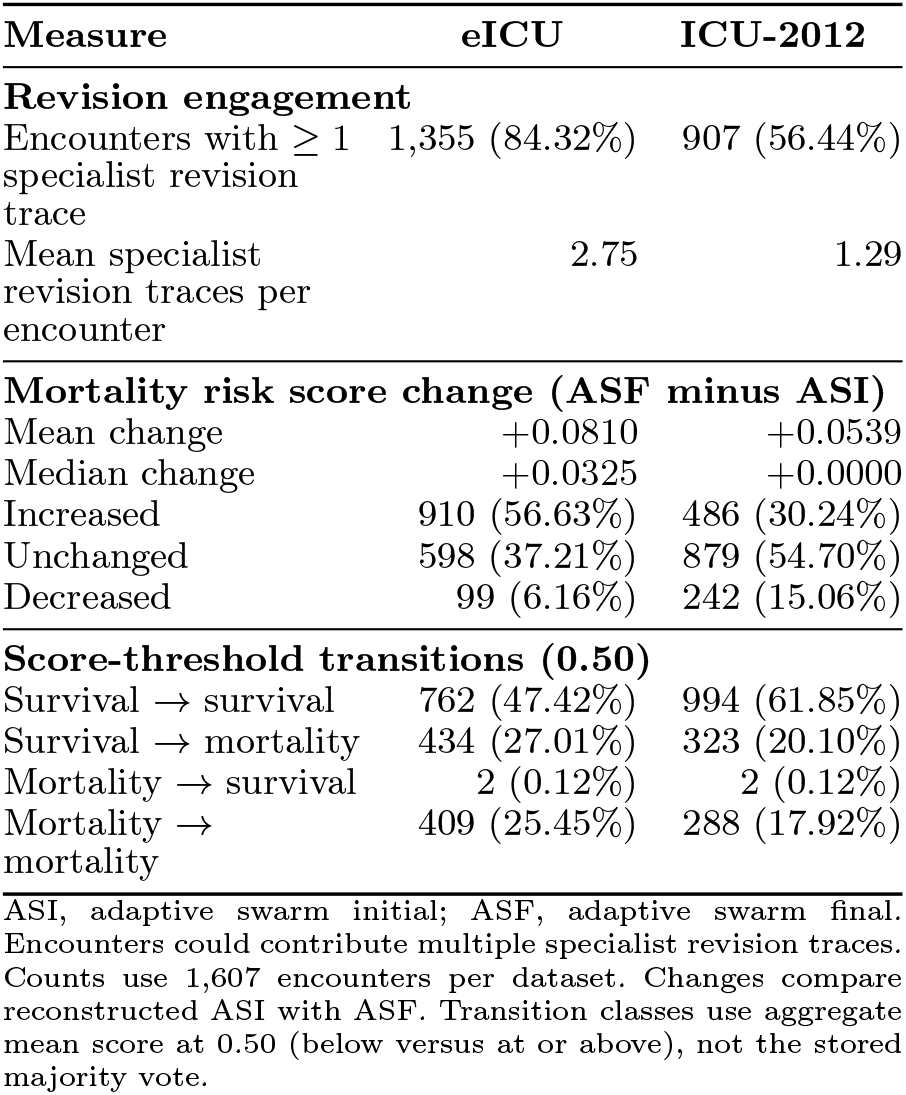
Adaptive-revision engagement, mortality risk score change, and score-threshold transitions.

**Fig. 2.**
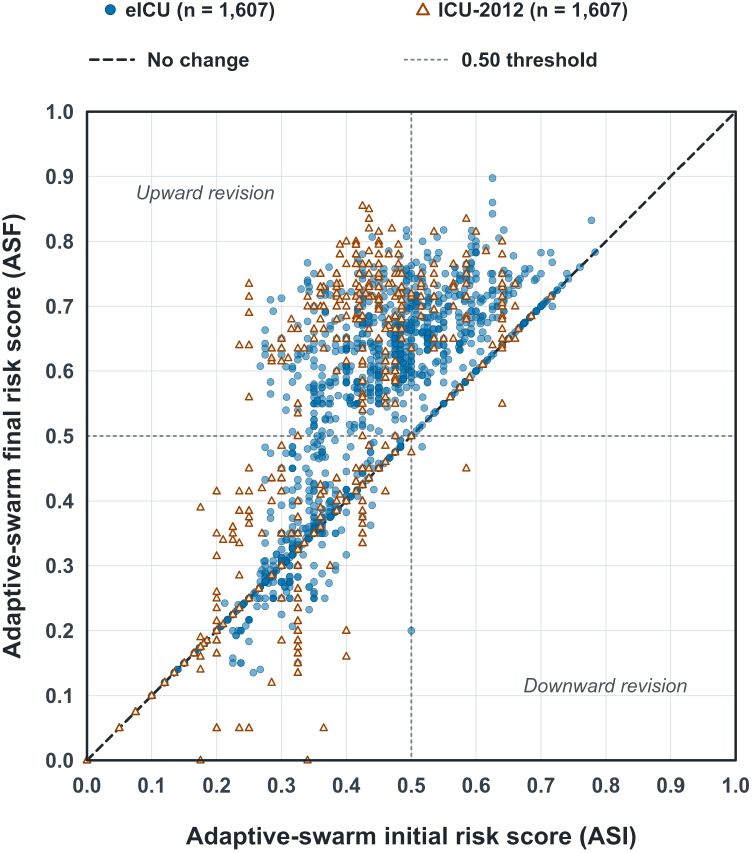
Encounter-level ASI-to-ASF mortality risk score drift. Blue filled circles show eICU and orange open triangles show ICU-2012. The diagonal marks no change and dotted lines the 0.50 analytic threshold; upper-left points crossed from the below-threshold to the at-or-above-threshold mortality classification.

Revision increased sensitivity, negative predictive value (NPV), and score-threshold mortality classification prevalence while reducing specificity and secondary accuracy in both datasets (Fig. 3). Brier scores worsened, while AUROC and AUPRC changes included zero. Balanced accuracy was unchanged and F1 decreased on eICU; both increased on ICU-2012. Positive predictive value (PPV) decreased on eICU, and PPV and MCC changes were uncertain otherwise.

**Fig. 3.**
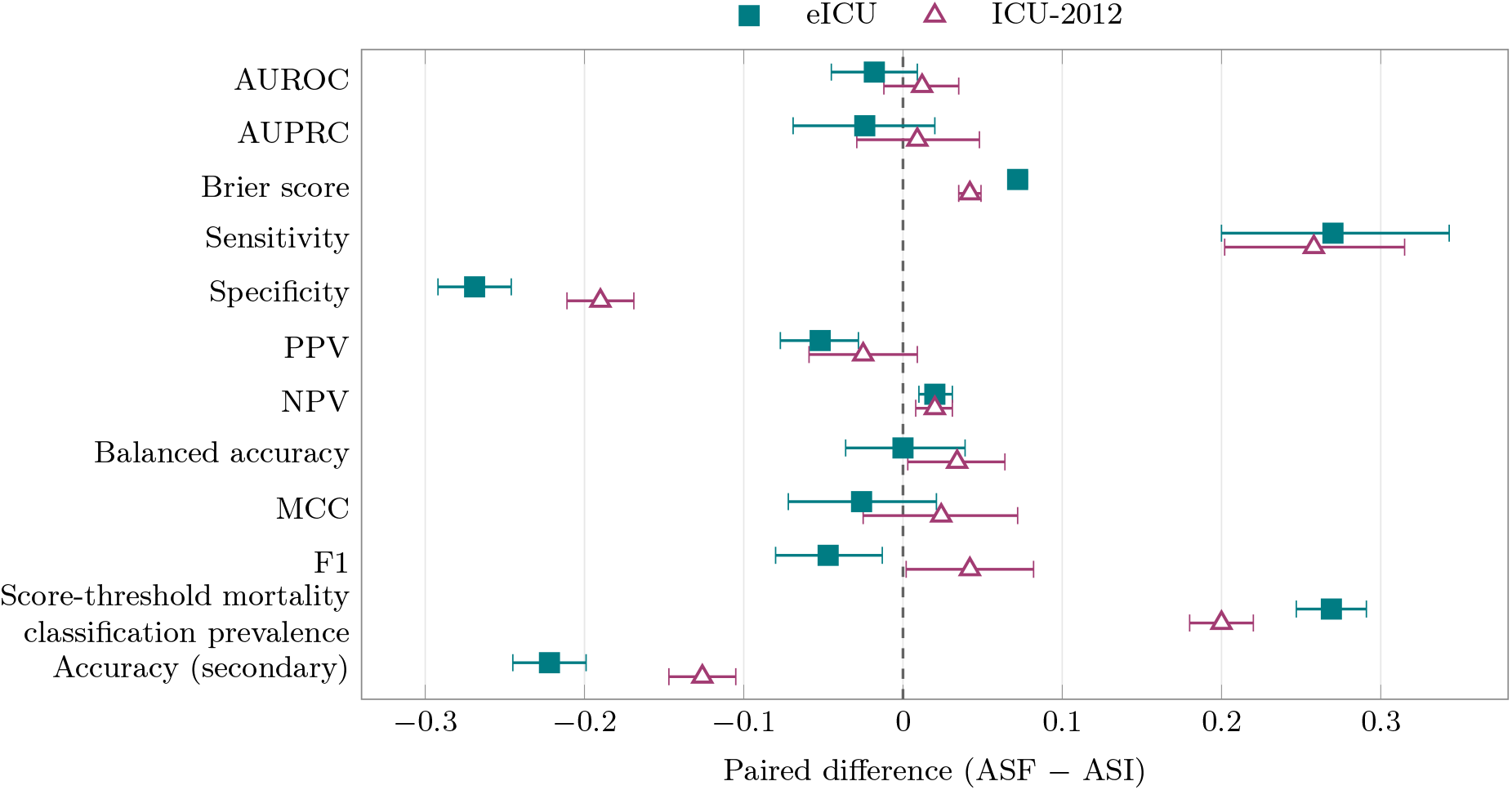
Paired changes after adaptive revision. Points show adaptive swarm final (ASF) minus adaptive swarm initial (ASI) estimates for 1,607 encounters per dataset; horizontal lines show hospital-stay-clustered 95% bootstrap confidence intervals. Positive values indicate increases rather than uniform improvement: lower Brier scores are preferable, the prevalence metric has no inherently favourable direction, and accuracy was secondary. PPV, positive predictive value; NPV, negative predictive value; MCC, Matthews correlation coefficient.

### 3.3 Evidence Use and Explanation Indicators

The independent end-to-end architecture benchmark showed mixed explanation indicators (Fig. 4). ASF used more supporting evidence and covered more specialist-cited evidence, but acknowledged counterevidence less often and produced more unsupported-claim flags. Citation validity was nearly complete and no explanation-validation failures occurred. Because specialist calls and final-explanation contexts differed, these contrasts do not estimate revision alone.

**Fig. 4.**
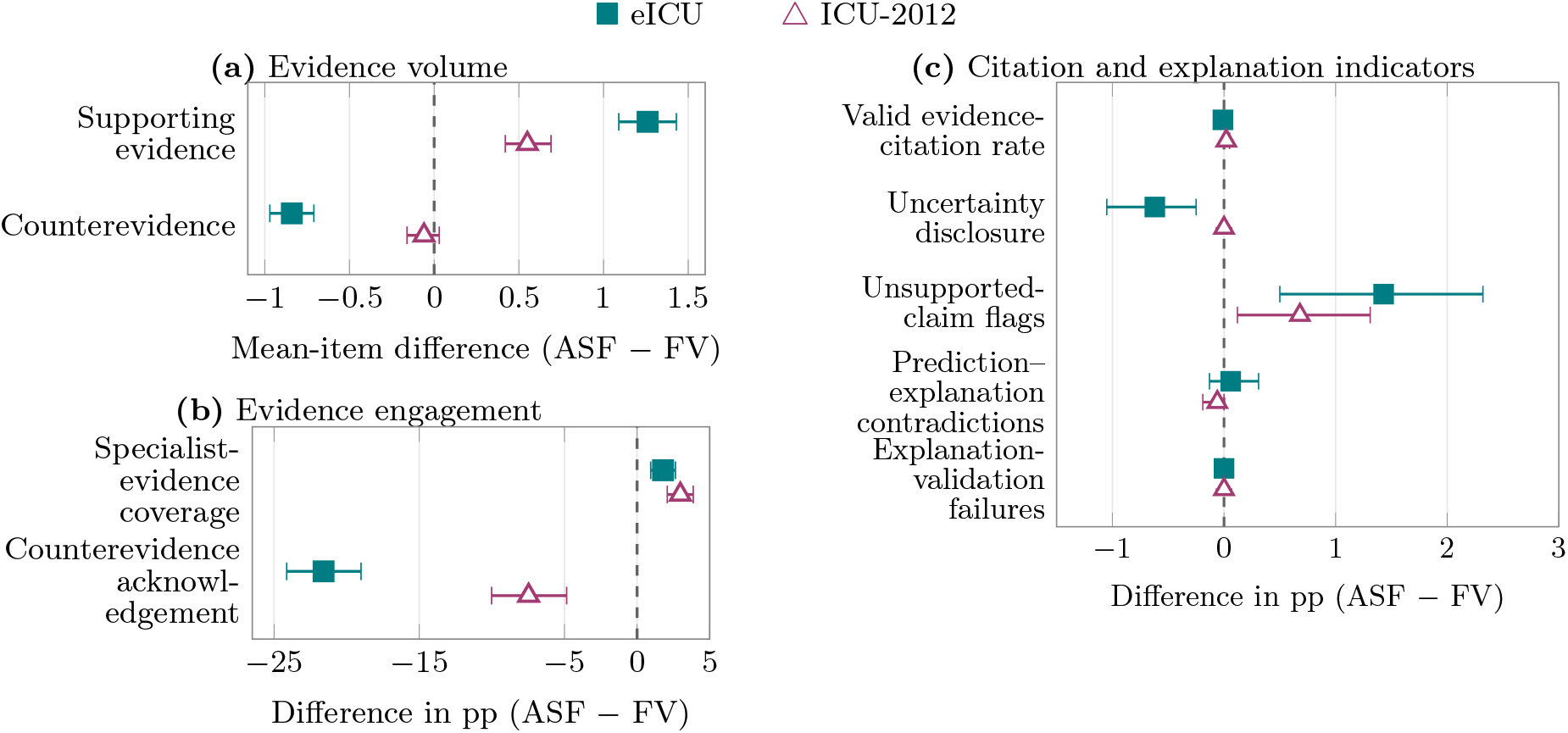
Matched-encounter ASF-minus-FV differences for independently executed architectures. Points show estimates and hospital-stay-clustered 95% bootstrap confidence intervals. Panel (a) uses items per explanation; panels (b) and (c) use percentage points (pp). Positive values are not necessarily favourable. These artifact-level contrasts do not isolate revision and were not human-validated.

### 3.4 Auditability

All final records met the reconstruction-completeness criterion, but ASF had more audit warnings, invalid-output fallbacks, and unsupported-claim warnings (Table 3). Abstentions were more frequent with ASF on eICU and universal in ICU-2012 because two domains were unavailable. These are architecture-level differences, not isolated revision effects.

**Table 3.** Artifact-level auditability and process-integrity indicators.

| Metric | ASF | FV | ASF minus FV (95% CI) |
| --- | --- | --- | --- |
| **eICU** |  |  |  |
| Records meeting reconstruction-completeness criterion | 1,607 / 1,607 (100.00%) | 1,607 / 1,607 (100.00%) | +0.00 pp (+0.00 to +0.00) |
| Encounters with audit warnings | 1,008 / 1,607 (62.73%) | 834 / 1,607 (51.90%) | +10.83 pp (+8.46 to +13.21) |
| Encounters with invalid-output fallbacks | 134 / 1,607 (8.34%) | 81 / 1,607 (5.04%) | +3.30 pp (+1.78 to +4.87) |
| Encounters with unsupported-claim warnings | 306 / 1,607 (19.04%) | 58 / 1,607 (3.61%) | +15.43 pp (+13.43 to +17.41) |
| Encounters with abstentions | 677 / 1,607 (42.13%) | 619 / 1,607 (38.52%) | +3.61 pp (+1.99 to +5.25) |
| **ICU-2012** |  |  |  |
| Records meeting reconstruction-completeness criterion | 1,607 / 1,607 (100.00%) | 1,607 / 1,607 (100.00%) | +0.00 pp (+0.00 to +0.00) |
| Encounters with audit warnings | 897 / 1,607 (55.82%) | 826 / 1,607 (51.40%) | +4.42 pp (+2.18 to +6.78) |
| Encounters with invalid-output fallbacks | 268 / 1,607 (16.68%) | 190 / 1,607 (11.82%) | +4.85 pp (+2.68 to +7.03) |
| Encounters with unsupported-claim warnings | 46 / 1,607 (2.86%) | 14 / 1,607 (0.87%) | +1.99 pp (+1.06 to +2.92) |
| Encounters with abstentions | 1,607 / 1,607 (100.00%) | 1,607 / 1,607 (100.00%) | +0.00 pp (+0.00 to +0.00) |

ASF, adaptive swarm final; FV, fixed voting; CI, confidence interval; pp, percentage points. ASF and FV were independently executed; matching was by encounter, not shared call. Completeness required latest specialist outputs, aggregation rule, final explanation, and audit-warning field. Lower warning and fallback rates are preferable; no composite score was calculated.

### 3.5 Contextual Predictive Performance

Predictive performance was contextual. ASI-to-ASF changes appear in Fig. 3; Table 4 benchmarks the independent final architectures. ASF had no clear AUROC advantage over FV. At the 0.50 score threshold it classified mortality more often, increasing sensitivity and reducing specificity, and had higher Brier scores. These end-to-end differences are not attributable to revision alone.

**Table 4.**
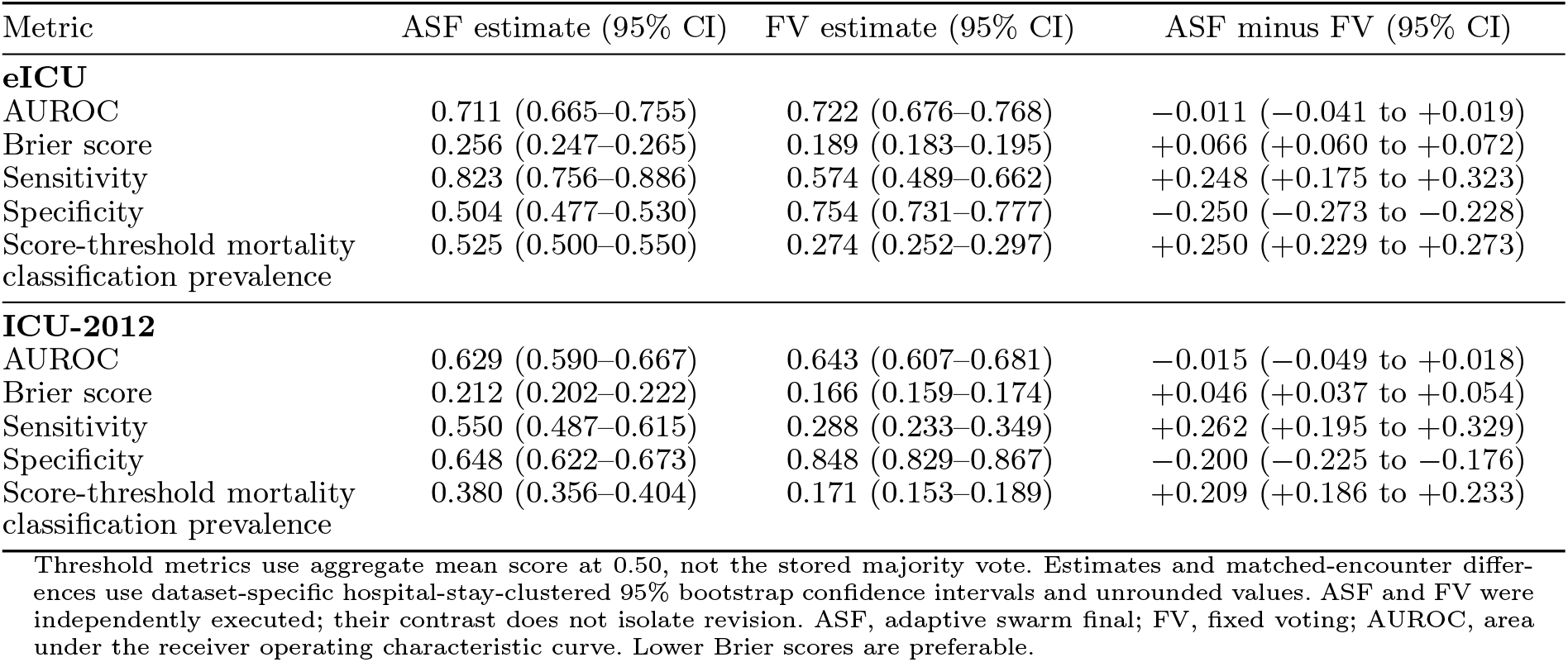
Selected contextual predictive performance of the final adaptive-swarm and fixed-voting outputs.

### 3.6 Computational Burden

ASF required more computation than FV. On eICU, ASF used a mean of 8.96 model calls per encounter versus 5.37 for FV (1.67 times as many) and required 101.43 versus 51.50 seconds per encounter (1.97 times the runtime). On ICU-2012, the corresponding values were 6.58 versus 5.06 calls (1.30 times as many) and 43.04 versus 28.35 seconds (1.52 times the runtime). Systematic ICU-2012 abstention affects cross-dataset comparisons; ASF–FV runtime differences compare the complete executions rather than revision alone.

## 4 Discussion

Within adaptive runs, revision changed operating behaviour more than risk-score ranking. Of 761 aggregate-score crossings at 0.50, 757 moved toward mortality, increasing sensitivity while reducing specificity and worsening Brier scores; AUROC and AUPRC showed no clear improvement. These score-threshold classes were not the stored majority-vote decisions. Effects varied by dataset and active specialist set, with two domains systematically absent in ICU-2012. Increased sensitivity from classifying more encounters as mortality is not by itself better prediction or evidence of clinical benefit [28, 34, 38].

In the independent architecture benchmark, ASF explanations used more supporting evidence but acknowledged counterevidence less often and had more unsupported-claim flags. These differences cannot be attributed to revision because specialist calls were separate and only the ASF explanation prompt received revision history. The pattern nevertheless cautions that richer artifacts can remain selectively incomplete [7, 11, 22, 31, 33]. A controlled revision study should reuse initial specialist outputs and hold the final-explanation prompt constant. Revision prompts should also require explicit handling of contradictory evidence and unresolved disagreement [7, 18].

All final records met the reconstruction-completeness criterion, indicating artifact availability rather than reliability. ASF also produced more warnings and fallbacks and required about twice the FV runtime on eICU and 1.5 times on ICU-2012. The additional stages may create both greater process visibility and more opportunities for detectable failure, but the architecture comparison cannot isolate revision.

Adaptive revision should therefore be treated as a configurable intervention, not a general performance upgrade. Future studies should prespecify triggers, validate thresholds externally, use common-start controls, and have clinicians evaluate explanations and handling of noisy inputs. Revision traces may support oversight but cannot substitute for validation, clinical utility, or accountability [9, 17, 19, 35].

## 5 Limitations

This retrospective study examined saved model outputs from two fixed cohorts and was not designed to assess prospective deployment, effects on clinician decisions, or patient outcomes; those questions require evaluation in the intended setting [17, 20, 28]. The clustered bootstrap accounted for repeated eICU encounters, but the confidence intervals describe uncertainty within the two evaluated cohorts rather than performance in new hospitals or populations. ICU-2012 records were not longitudinally linked, so each encounter was treated as its own bootstrap cluster; clustered resampling was therefore equivalent to encounter-level resampling. Cross-dataset comparisons were descriptive because the cohorts differed in mortality prevalence, clustering, available evidence, and specialist participation. Such differences can affect transportability through shifts in populations, clinical data-generating processes, and site-specific acquisition or workflow patterns [12, 16, 21]. In particular, two specialists abstained for every ICU-2012 encounter because their clinical domains were unavailable, reducing the active agent set and affecting revision engagement and computational burden.

Scores were uncalibrated, and 0.50 served only as a consistent analytic operating point. Score-threshold classes were distinct from stored majority-vote decisions; neither was intended for clinical use [29, 34].

ASF and FV were independently executed. Their contrasts include separate specialist responses and explanation contexts and therefore cannot identify a revision effect. The bootstrap captures cohort sampling conditional on the saved runs, not between-run LLM variability; causal comparison requires common-start repeated runs.

Plausibility filtering was intentionally omitted so behavior under noisy inputs remained observable. Predictive estimates are therefore not optimized clinical performance, and responses to individual artifacts were not clinically adjudicated.

Explanation and auditability measures were automated artifact properties, not human assessments. Faithfulness, usefulness, user impact, and decision validity require separate evaluation [2, 10, 19, 32].

Findings may not generalize beyond the evaluated model, prompts, rules, and executions [7, 18, 31]. Unadjusted confidence intervals across multiple outcomes are exploratory. Fairness and subgroup performance were not evaluated, so no subgroup-equity conclusions should be drawn.

## 6 Conclusion

Within adaptive runs, revision shifted aggregate scores across the 0.50 threshold toward mortality without clear discrimination improvement. Independently executed ASF outputs used more supporting evidence than FV but acknowledged counterevidence less often, generated more warnings, and required more computation. Because specialist calls and explanation contexts were not shared, these architecture contrasts do not isolate revision. Future common-start, repeated-run studies should assess aggregation, counterevidence handling, and clinician review of inputs and traces [2, 10, 19, 32].

## Statements and Declarations

### Author Contributions

C.A. conceived the study, designed the methodology, implemented the software, conducted the experiments, analyzed the results, prepared the figures and tables, and wrote and revised the manuscript.

### Ethics Approval

This study used only retrospective, de-identified records from the publicly accessible eICU Collaborative Research Database Demo and PhysioNet/Computing in Cardiology Challenge 2012 datasets, involved no direct participant contact or intervention, and made no attempt to re-identify individuals. The source datasets were released under their original institutional governance and de-identification procedures.

### Data Availability

The source data are publicly available from PhysioNet: the eICU Collaborative Research Database Demo, version 2.0.1, at https://physionet.org/content/eicu-crd-demo/2.0.1/, and the PhysioNet/Computing in Cardiology Challenge 2012, version 1.0.0, at https://physionet.org/content/challenge-2012/1.0.0/. Access and reuse are subject to the terms specified by PhysioNet and the respective dataset licences. No patient-level source data are redistributed in the study repository.

### Code Availability

Source code, prompts, configuration files, and analysis scripts are available at https://github.com/CallumGA/swarm-ehr. The repository documents the software dependencies and procedures used for cohort processing, architecture execution, and analysis.

### Study Protocol and Registration

No separate study protocol was prepared, and the study was not prospectively registered.

### Patient and Public Involvement

Patients and members of the public were not involved in the design, conduct, reporting, interpretation, or dissemination planning of this retrospective methodological study.

## References

[1] Anderson C (2026) Agent role structure and operating characteristics in large language model clinical classification: A comparative study of specialist and deliberative multi-agent protocols. Informatics in Medicine Unlocked 64:101783. 10.1016/j.imu.2026.101783, URL https://doi.org/10.1016/j.imu.2026.101783

[2] Awasthi R, Bhattad A, Ramachandran SP, et al (2025) Human evaluation of large language models in healthcare: Gaps, challenges, and the need for standardization. npj Health Systems 2:40. 10.1038/s44401-025-00043-2, URL https://www.nature.com/articles/s44401-025-00043-2

[3] Bedi S, Liu Y, Orr-Ewing L, et al (2025) Testing and evaluation of health care applications of large language models: A systematic review. JAMA 333(4):319–328. https://doi.org/10.1001/jama.2024.21700, URL 10.1001/jama.2024.21700

[4] Chen X, Yi H, You M, et al (2025) Enhancing diagnostic capability with multi-agents conversational large language models. npj Digital Medicine 8(1):159. 10.1038/s41746-025-01550-0, URL https://doi.org/10.1038/s41746-025-01550-0

[5] Collins GS, Moons KGM, Dhiman P, et al (2024) TRIPOD+AI statement: Updated guidance for reporting clinical prediction models that use regression or machine learning methods. BMJ 385:e078378. 10.1136/bmj-2023-078378, URL https://doi.org/10.1136/bmj-2023-078378

[6] Contreras M, Silva B, Shickel B, et al (2025) Real-time prediction of intensive care unit patient acuity and therapy requirements using state-space modelling. Nature Communications 16:7315. 10.1038/s41467-025-62121-1, URL https://www.nature.com/articles/s41467-025-62121-1

[7] Estornell A, Liu Y (2024) Multi-LLM debate: Framework, principals, and interventions. In: Advances in Neural Information Processing Systems, 10.52202/079017-0911, URL https://proceedings.neurips.cc/paper_files/paper/2024/hash/32e07a110c6c6acf1afbf2bf82b614ad-Abstract-Conference.html

[8] Gaber F, Shaik M, Allega F, et al (2025) Evaluating large language model workflows in clinical decision support for triage and referral and diagnosis. npj Digital Medicine 8:263. 10.1038/s41746-025-01684-1, URL https://www.nature.com/articles/s41746-025-01684-1

[9] Gallifant J, Afshar M, Ameen S, et al (2025) The TRIPOD-LLM reporting guideline for studies using large language models. Nature Medicine 31(1):60–69. 10.1038/s41591-024-03425-5, URL https://www.nature.com/articles/s41591-024-03425-5

[10] Ghanvatkar S, Rajan V (2024) Evaluating explanations from AI algorithms for clinical decision-making: A social science-based approach. IEEE Journal of Biomedical and Health Informatics 28(7):4269–4280. 10.1109/JBHI.2024.3393719, URL https://doi.org/10.1109/JBHI.2024.3393719

[11] Ghassemi M, Oakden-Rayner L, Beam AL (2021) The false hope of current approaches to explainable artificial intelligence in health care. The Lancet Digital Health 3(11):e745–e750. 10.1016/S2589-7500(21)00208-9, URL https://www.sciencedirect.com/science/article/pii/S2589750021002089

[12] Goetz L, Seedat N, Vandersluis R, et al (2024) Generalization—a key challenge for responsible AI in patient-facing clinical applications. npj Digital Medicine 7:126. 10.1038/s41746-024-01127-3, URL https://www.nature.com/articles/s41746-024-01127-3

[13] Hegselmann S, Shen Z, Gierse F, et al (2024) A data-centric approach to generate faithful and high quality patient summaries with large language models. In: Proceedings of the Fifth Conference on Health, Inference, and Learning, Proceedings of Machine Learning Research, vol 248. PMLR, pp 339–379, URL https://proceedings.mlr.press/v248/hegselmann24a.html

[14] Ke Y, Yang R, Lie SA, et al (2024) Mitigating cognitive biases in clinical decision-making through multi-agent conversations using large language models: Simulation study. Journal of Medical Internet Research 26:e59439. 10.2196/59439, URL https://www.jmir.org/2024/1/e59439/

[15] Kim Y, Park C, Jeong H, et al (2024) MDA-gents: An adaptive collaboration of LLMs for medical decision-making. In: Advances in Neural Information Processing Systems, 10.52202/079017-2522, URL https://proceedings.neurips.cc/paper_files/paper/2024/hash/90d1fc07f46e31387978b88e7e057a31-Abstract-Conference.html

[16] Lasko TA, Strobl EV, Stead WW (2024) Why do probabilistic clinical models fail to transport between sites. npj Digital Medicine 7:53. 10.1038/s41746-024-01037-4, URL https://www.nature.com/articles/s41746-024-01037-4

[17] Lekadir K, Frangi AF, Porras AR, et al (2025) FUTURE-AI: International consensus guideline for trustworthy and deployable artificial intelligence in healthcare. BMJ 388:e081554. 10.1136/bmj-2024-081554, URL https://www.bmj.com/content/388/bmj-2024-081554

[18] Li J, Deng Y, Sun Q, et al (2025) Benchmarking large language models in evidence-based medicine. IEEE Journal of Biomedical and Health Informatics 29(9):6143–6156. 10.1109/JBHI.2024.3483816, URL https://doi.org/10.1109/JBHI.2024.3483816

[19] Livingston L, Featherstone-Uwague A, Barry A, et al (2025) Reproducible generative artificial intelligence evaluation for health care: A clinician-in-the-loop approach. JAMIA Open 8(3):ooaf054. 10.1093/jamiaopen/ooaf054, URL https://academic.oup.com/jamiaopen/article/8/3/ooaf054/8163901

[20] Moons KGM, Damen JAA, Kaul T, et al (2025) PROBAST+AI: An updated quality, risk of bias, and applicability assessment tool for prediction models using regression or artificial intelligence methods. BMJ 388:e082505. 10.1136/bmj-2024-082505, URL https://www.bmj.com/content/388/bmj-2024-082505

[21] Ong Ly C, Unnikrishnan B, Tadic T, et al (2024) Shortcut learning in medical AI hinders generalization: Method for estimating AI model generalization without external data. npj Digital Medicine 7:124. 10.1038/s41746-024-01118-4, URL https://www.nature.com/articles/s41746-024-01118-4

[22] Paul D, West R, Bosselut A, et al (2024) Making reasoning matter: Measuring and improving faithfulness of chain-of-thought reasoning. In: Findings of the Association for Computational Linguistics: EMNLP 2024. Association for Computational Linguistics, pp 15012–15032, 10.18653/v1/2024.findings-emnlp.882, URL https://aclanthology.org/2024.findings-emnlp.882/

[23] Pollard T, Johnson A, Raffa J, et al (2019) eICU collaborative research database. PhysioNet 10.13026/C2WM1R, URL https://doi.org/10.13026/C2WM1R, version 2.0

[24] Pollard T, Moody BE, Lehman LwH, et al (2026) PhysioNet as a global platform for biomedical research. Nature Health 1:792–795. 10.1038/s44360-026-00096-z, URL https://doi.org/10.1038/s44360-026-00096-z

[25] Pollard TJ, Johnson AEW, Raffa JD, et al (2018) The eICU collaborative research database, a freely available multi-center database for critical care research. Scientific Data 5:180178. 10.1038/sdata.2018.178, URL https://doi.org/10.1038/sdata.2018.178

[26] Qwen Team (2026) Qwen3.5: Towards native multimodal agents. URL https://qwen.ai/blog?id=qwen3.5

[27] Rezgui K (2024) Large language models for healthcare: Applications, models, datasets, and challenges. In: 2024 10th International Conference on Control, Decision and Information Technologies (CoDIT). IEEE, pp 2366–2371, 10.1109/CoDIT62066.2024.10708253, URL https://doi.org/10.1109/CoDIT62066.2024.10708253

[28] Riley RD, Archer L, Snell KIE, et al (2024) Evaluation of clinical prediction models (part 2): How to undertake an external validation study. BMJ 384:e074820. 10.1136/bmj-2023-074820, URL https://www.bmj.com/content/384/bmj-2023-074820

[29] Savage T, Wang J, Gallo RJ, et al (2025) Large language model uncertainty proxies: Discrimination and calibration for medical diagnosis and treatment. Journal of the American Medical Informatics Association 32(1):139–149. 10.1093/jamia/ocae254, URL https://academic.oup.com/jamia/article/32/1/139/7819854

[30] Silva I, Moody G, Scott DJ, et al (2012) Predicting in-hospital mortality of ICU patients: The PhysioNet/computing in cardiology challenge 2012. In: Computing in Cardiology 2012, pp 245–248, URL https://physionet.org/files/challenge-2012/1.0.0/papers/0245.pdf

[31] Smit AP, Grinsztajn N, Duckworth P, et al (2024) Should we be going MAD? A look at multi-agent debate strategies for LLMs. In: Proceedings of the 41st International Conference on Machine Learning, Proceedings of Machine Learning Research, vol 235. PMLR, pp 45883–45905, URL https://proceedings.mlr.press/v235/smit24a.html

[32] Tam TYC, Sivarajkumar S, Kapoor S, et al (2024) A framework for human evaluation of large language models in healthcare derived from literature review. npj Digital Medicine 7:258. 10.1038/s41746-024-01258-7, URL https://www.nature.com/articles/s41746-024-01258-7

[33] Turpin M, Michael J, Perez E, et al (2023) Language models don’t always say what they think: Unfaithful explanations in chain-of-thought prompting. In: Advances in Neural Information Processing Systems, 10.52202/075280-3275, URL https://proceedings.neurips.cc/paper_files/paper/2023/hash/ed3fea9033a80fea1376299fa7863f4a-Abstract-Conference.html

[34] Van Calster B, Collins GS, Vickers AJ, et al (2025) Evaluation of performance measures in predictive artificial intelligence models to support medical decisions: Overview and guidance. The Lancet Digital Health 7(12):100916. 10.1016/j.landig.2025.100916, URL https://www.sciencedirect.com/science/article/pii/S2589750025000986

[35] Vasey B, Nagendran M, Campbell B, et al (2022) Reporting guideline for the early stage clinical evaluation of decision support systems driven by artificial intelligence: DECIDE-AI. BMJ 377:e070904. 10.1136/bmj-2022-070904, URL https://www.bmj.com/content/377/bmj-2022-070904

[36] Wang X, Yang CC (2026) Medihive: A decentralized agent collective for medical reasoning. Journal of Healthcare Informatics Research 10.1007/s41666-026-00239-7, URL https://doi.org/10.1007/s41666-026-00239-7

[37] Wang Z, Zhu Y, Zhao H, et al (2025) ColaCare: Enhancing electronic health record modeling through large language model-driven multi-agent collaboration. In: WWW ‘25: Proceedings of the ACM on Web Conference 2025. Association for Computing Machinery, pp 2250–2261, 10.1145/3696410.3714877, URL https://dl.acm.org/doi/10.1145/3696410.3714877

[38] Wells BJ, Nguyen HM, McWilliams A, et al (2025) A practical framework for appropriate implementation and review of artificial intelligence (FAIR-AI) in healthcare. npj Digital Medicine 8:514. 10.1038/s41746-025-01900-y, URL https://www.nature.com/articles/s41746-025-01900-y

[39] Xie P, Hu Y, Li J, et al (2025) Unlocking the potential of real-time ICU mortality prediction: Redefining risk assessment with continuous data recovery. npj Digital Medicine 8:733. 10.1038/s41746-025-02114-y, URL https://www.nature.com/articles/s41746-025-02114-y

